# Why a Globally Fair COVID-19 Vaccination? An Analysis based on Agent-Based Simulation

**DOI:** 10.1101/2021.10.03.21264494

**Authors:** Kashif Zia

## Abstract

In this paper, an Agent-Based Model (ABM) is proposed to evaluate the impact of COVID-19 vaccination drive in different settings. The main focus is to evaluate the counter-effectiveness of disparity in vaccination drive among different regions / countries. The model proposed is simple yet novel in the sense that it captures the spatial transmission-induced activity into consideration, through which we are able to relate transmission model to the mutated variations of the virus. Some important what-if questions are asked in terms of number of deaths, and time required and percentage of population needed to be vaccinated before the pandemic is eradicate. The simulation results have revealed that it is necessary to maintain a global (rather than regional or country oriented) vaccination drive in case of a new pandemic or continual efforts against COVID-19.

## 1. Introduction

After the successful provisioning of COVID-19 vaccination, there has been efforts to achieve a global herd immunity as soon as possible [1]. However, disparity in vaccination drive among different countries can be counter-productive. A general evidence of health disparity in COVID-19 treatments if given in [2]. There is a need to analyze the effects of vaccine provisioning disparity at a global scale [3].

A very few countries are involved in the process of vaccine production. Obviously, these countries would vaccinate their own population first before the vaccine is available to other countries. However, particularly in case of such a complex global system, the intuitive logic, quite obvious in a situation may not be the optimal choice. Through this paper, we provide an evidence to establish it in the context of COVID-19 vaccination drive.

In this paper, an Agent-Based Model (ABM) is proposed to evaluate the impact of COVID-19 vaccination drive in different settings. The simulation results have revealed that it is necessary to maintain a globally “fair” [4] vaccination drive in case of a new pandemic or continual efforts against COVID-19. Here fair means a vaccination drive which is not self-centered (focused on developed countries producing the vaccine), but global (equally distributed right from day 1) across all the regions / countries.

A simplistic model of virus transmission is used consisting of minimal states of susceptible, vaccinated, infected and recovered. A moving agent in one of these states is tightly bound to the underlying space, where the space is divided into regions to evaluate the region-based vs. global vaccination drive. Additionally, the virus gets mutated, where the extent of mutation is directly related to spatial activity representing the transmissions. And the inactivity is directly proportional to the mutated variant at a location.

Already, a few agent-based models concerning the vaccination efficiency have been proposed. Authors in [5] have proposed vaccination strategies with a delayed second dose. They have compared different vaccination products and provided a projected number of infections, serious cases and deaths. Authors in [6] proposed a mathematical model to estimate the impact on mortality and total infections of completely lifting the COVID-19 restrictions. A qualitative study on who should be prioritised for COVID-19 vaccination is given in [7]. Another focused attempt is a simulation study to estimate the future infections rates among the children with vaccination [8].

More closer to our paper is [9], in which the authors emphasize on accelerating the vaccination drive to mitigate high transmissibility resulting in more deadly variants. However, the model proposed is population based without spatial (regional / country-wise) variations. Another similar model is presented by the authors in [10]. But the system dynamic model again does not cater for spatial considerations and mobility. Another sound research is published in [11], in which the authors combines the real data with the agent-based model to estimate the impact of lockdown and vaccination against COVID-19. But, this model is about one country only and does not take nonavailability of vaccine as an option.

As evidenced by the above, the model proposed in this paper is simple, but, novel in the sense that it captures the spatial virus transmission-related activity into consideration. This enable us to do inquiry about the effectiveness of the vaccination drive in different settings. In the next section, we have detailed the model, followed by simulation and results section. The last section provide a detailed outlook of this research in progress.

## 2. Model

The model represents an individual as a mobile agent, which resides on a static cell. The epidemic spread model is based on simple **susceptible**-**infected**-**recovered**/not recovered (SIR) model to keep focus on vaccination aspect. In addition to these three states, the fourth state of being **vaccinated** is added. A few agents in the population are made infected at the start of the simulation. Causally, a susceptible agent can directly be transited to infected state or it may transit to vaccinated state. An agent may also be transited to infected state from vaccinated state. An infected agent may be recovered of may not (die). Even a recovered agent can still be susceptible.

The simulation runs in iterations. At each time stamp, each of the agents make a causal change in its state depending upon its surrounding. The model is implemented through three sequential modules (procedures), namely transmission, recovery and vaccination, followed by running the module which let the agents move randomly.

### 2.1. Transmission

This procedure is executed by all the agents (randomly ordered) which are infected now. The procedure is dependent on the following variables:

- *transmission_rate* of agent **a** (one of the infected agent) pinned onto the location transmissibility. The location transmissibility depends on how transmission intensive the neighborhood of agent **a** is. This in turn is handled by cell (patch) variable *activity*, which is increased as soon as a transmission happens due to the resident agent. The sample activity maps comparing transmission activity of two cases (Figure 1a vs. Figure 1b).
- A neighboring agent **n** which is susceptible.
- Neighborhood-based variable *ratio_vaccinated*, which is the ratio of neighborhood agents of agent **n** that are in vaccinated state.

**Figure 1:**
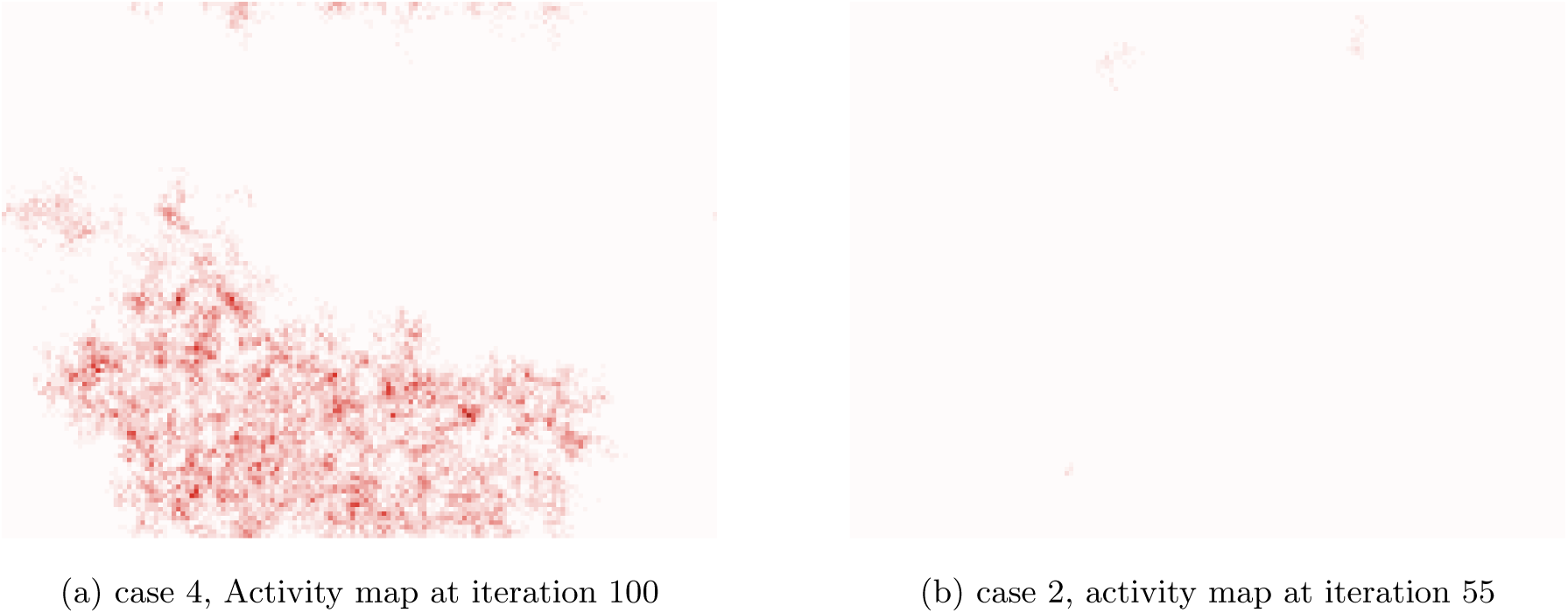
Virus spatial activity: case 4 and 2.

**Figure 2:**
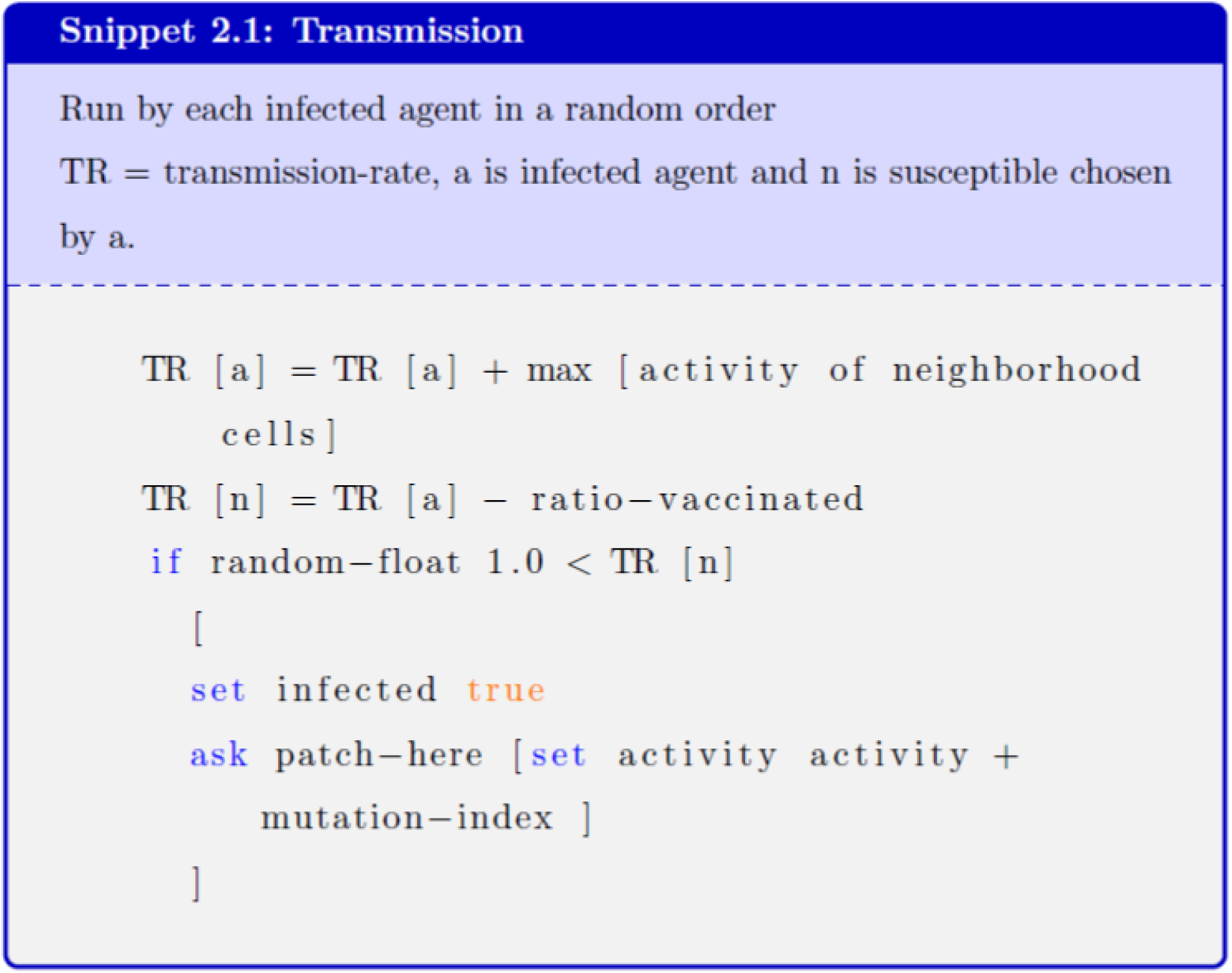
Transmission

**Figure 3:**
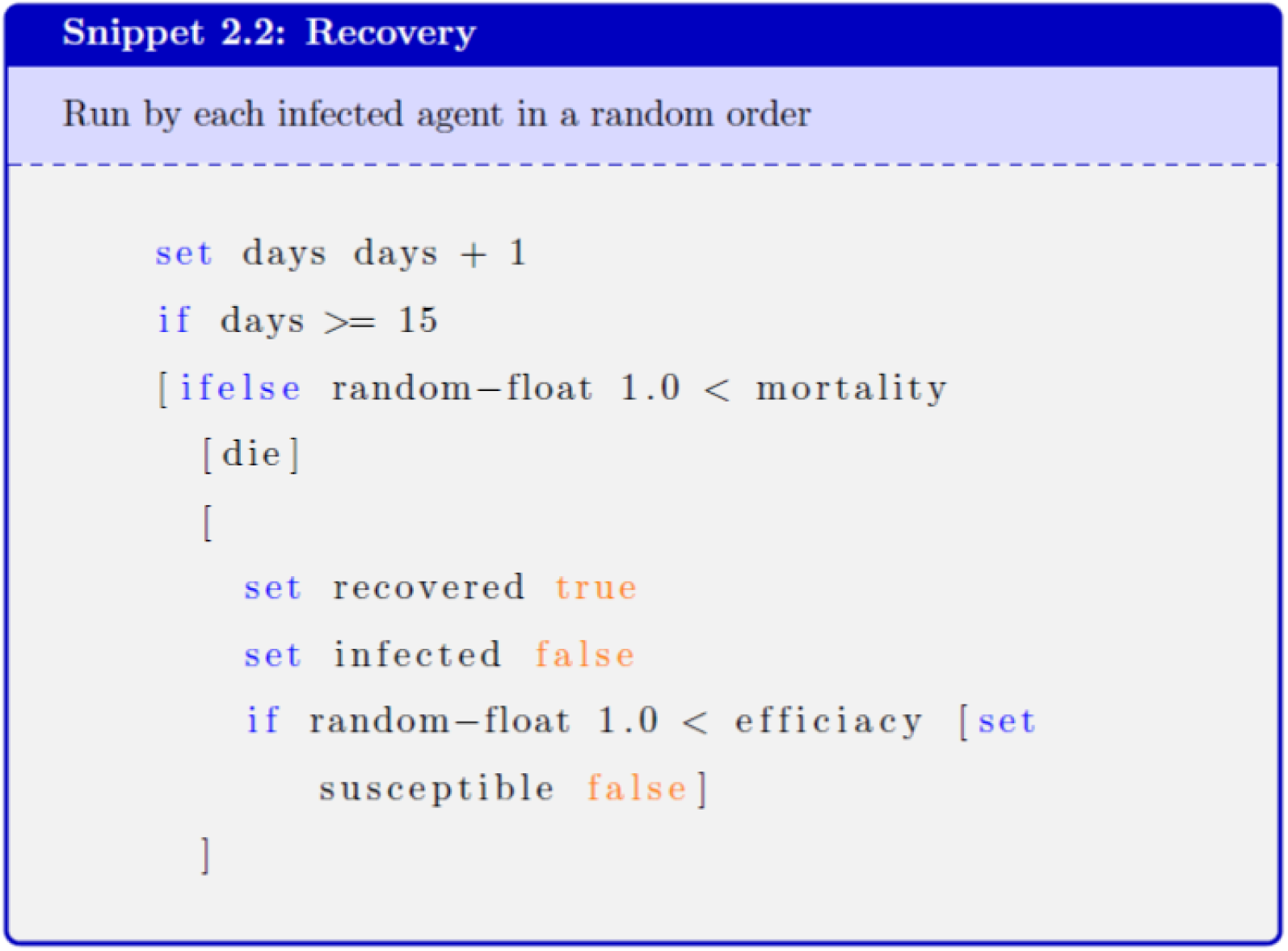
Recovery

**Figure 4:**
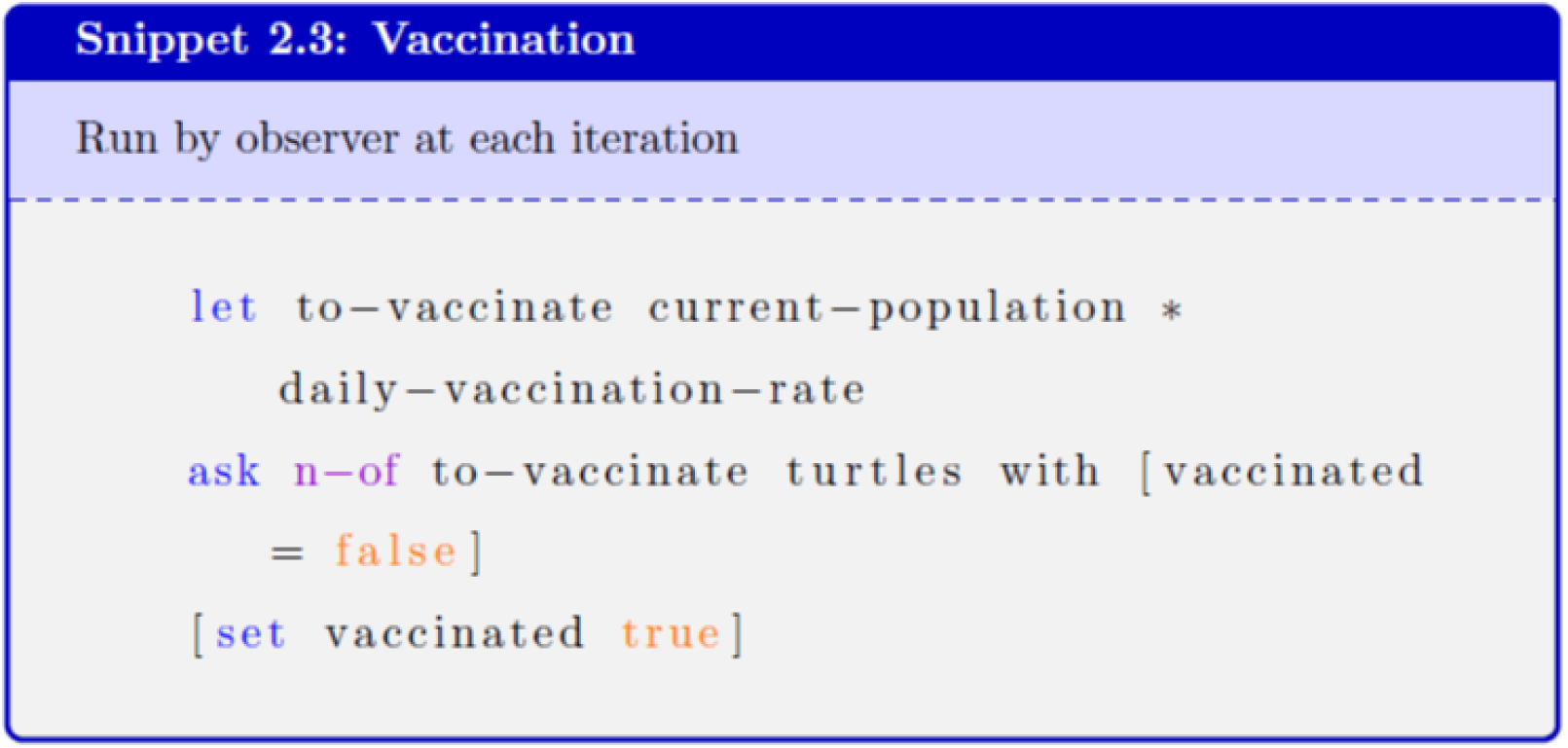
Vaccination

If an agent a has a susceptible agent n in its neighborhood, agent n becomes infected with a probability defined by transmission_rate (TR [n]) that is been encountered. TR [n] is equal to difference of transmission_rate of the source agent (TR [a]) and the ratio of agents already vaccinated (ratio_vaccinated) in the neighborhood of agent n. Hence, the intensity by which the infection is induced by a is depleted based on vaccination. TR [a] is incremented before, depending on how transmission active the neighborhood of a has been. If n is infected, the activity of underlying patch is also increased by a factor mutation_index, which represents how effectively mutation is changing the local transmission. The pseudo code given in Snippet 2.1 describes the transmission mechanism run by each infected agent:

### 2.2. Recovery

This procedure applies to those agents which are infected. Each of these agents either remains in the infected state, gets recovered, or dies, depending on the number of days it has been infected and chances (controlled by simulation parameters of mortality and efficacy). The self-explanatory pseudo code given as Snippet 2.2 describes the mechanism run by each infected agent:

### 2.3. Vaccination

According to a vaccination rate, the agents which are not yet vaccinated are vaccinated. This is a simple procedure just setting state of an agent to true. The self-explanatory pseudo code given as Snippet 2.3 describes the vaccination process:

## 3. Simulation

A cell space of size 160 *×* 120 is used as the simulation world, wrapped around horizontally as well as vertically. Therefore a population of a maximum of 19200 non-overlapping agents can be generated, each occupying a single patch. We opted to take 50% of the population (9600 agents). Each patch is first initialized with activity 0. As the simulation progresses, all the agents move on patches with a designated speed. They also change their health states (based on the model above), and may end up as dead agents, thus no more part of the simulation world.

In Table 1, the list of global simulation parameters are given. The other parameters such as transmission range (radius of neighborhood), mortality rate, and efficacy of vaccine are also used, however, these acquire static values of 1.5 patch distance, 0.2, and 0.8, respectively. Whereas, the last two parameters define the four cases we have considered. These cases are given in Table 2. **What do these cases mean?** Since, in the simulation, the vaccination happens from day 1, the vaccination rate equal to 0% means that the transmission of virus happens along with the vaccination drive. Obviously, this did not happen in case of COVID-19. Hence, 0% vaccination rate depicts a futuristic (ideal) situation in which vaccination is available before the virus outbreak happens. Whereas, a vaccination rate of 20% is about 20% population already vaccinated when the transmission starts. This can be equated to (probably) end of second global wave of COVID-19, when the active cases were quite low and a substantial population was vaccinated. Therefore, the prior represents an **ideal** and the later represents a **real** scenario (but an intermediate one).

**Table 1:** Global Simulation Parameters.

| Parameter | Description | Values |
| --- | --- | --- |
| Population_size | The total number of agents | Initialized with static value of 50% of the space |
| Transmission_rate | Transmission rate of the virus | Initialized with 0.50, where all the agents clone the same value initially, which may change later on as given in transmission procedure |
| Mutation_index | The factor by which Transmission_rate is increased | Initialized static value of 0.02 |
| Vaccination_rate | The percentage of people who are already vaccinated (at the start of the simulation) | Either 0% or 20% |
| blocked? | whether the space used is divided into blocks or not | Either true or false |

**Table 2:** Simulation cases.

| Case | blocked? | Vaccination_rate | scenario |
| --- | --- | --- | --- |
| case1 | false | 0 | global ideal |
| case2 | false | 20 | global real |
| case3 | true | 0 | self-centered ideal |
| case4 | true | 20 | self-centered real |

Since, the vaccination drive (as we have seen in case of COVID-19) was / has been country-centered, where the (mostly) developed countries vaccinated their own population first, followed by provision of the vaccine to other countries. We term this scenario as **self-centered**, and it is implemented in the simulation by introduction of regional blocks (blocks? = true). Therefore, some blocks have more vaccines in comparison to others. An alternate strategy would have been a globally balanced vaccination drive irrespective of country of manufacturing. Thus, that would have been a **global** drive, in which, all the blocks would have been the same (hence, block? = false).

In fact, there are 12 regional blocks as shown in Figure 5. Although, a clear differentiation is enough for modeling purposes, however, the blocks/regions are sorted from left to right and from bottom to top, in terms of availability of vaccine. So for example, region 1 is the worst, followed by region 2, 3, and so on. The last column of Table 2 summarizes the scenarios in terms of the above two dimensions.

**Figure 5:**
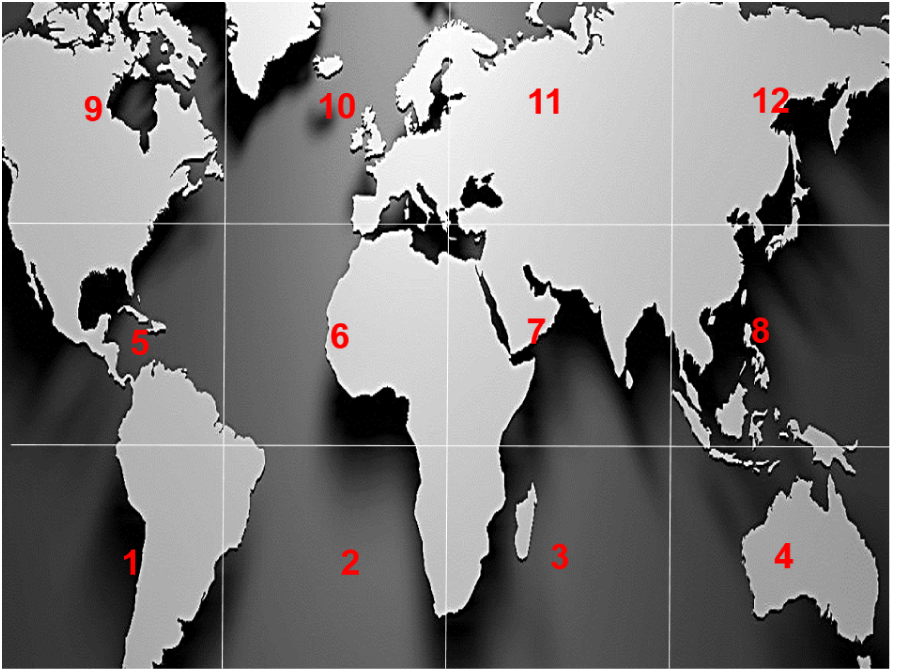
World arranged in vaccination blocks.

So the idea is to measure the performance of the vaccination drive in all four scenarios and do a comparative analysis. The simulations are average of sufficient number of runs. The results are compared based on the following outcomes. The results shown in graph in Figure 6 represent the number of agents in different states after the simulation ends (there is no more infected agent in the population). The graph in Figure 9 shows the number of people dead and time when the simulation ends, with particular focus on blocks situation.

**Figure 6:**
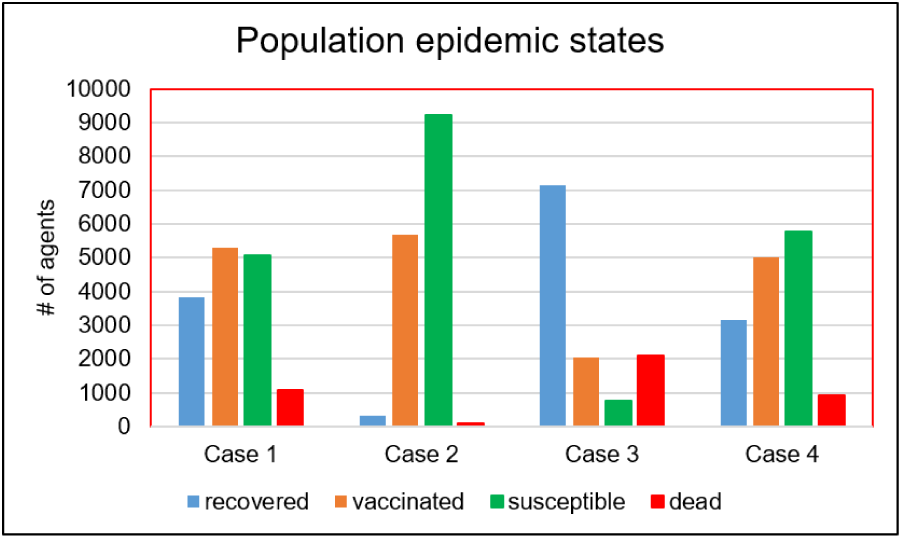
Epidemic states at the end of the simulation.

### states

- Starting with the self-centered real scenario (case 4), it can be seen that, we end up with almost 10% of the population dead, whereas, recovered, vaccinated and susceptible follow the same order. If we compare these results with global real scenario (case 2), the dead % drops down to less than 1 (see Figure 7). It should be noted that a very small fraction of infected agents (taken value of 0.005-0.010%) dispersed randomly across the space at the start of the simulation, does not really represents the reality. However, it can be considered as cancelling the fact that vaccinations are also done at random, not according to the severity of the regional infectiousness. Nevertheless, it does not take away the fact that a global vaccination drive would have helped a lot.
- For the ideal scenarios, case 3 represents the self-centered situation. The dead % increases to 22. Whereas, for a global situation (case 1), it could be reduced to 9.5%. However, case 3 is much more diversified in terms of distribution of deaths (see Figure 8).

**Figure 7:**
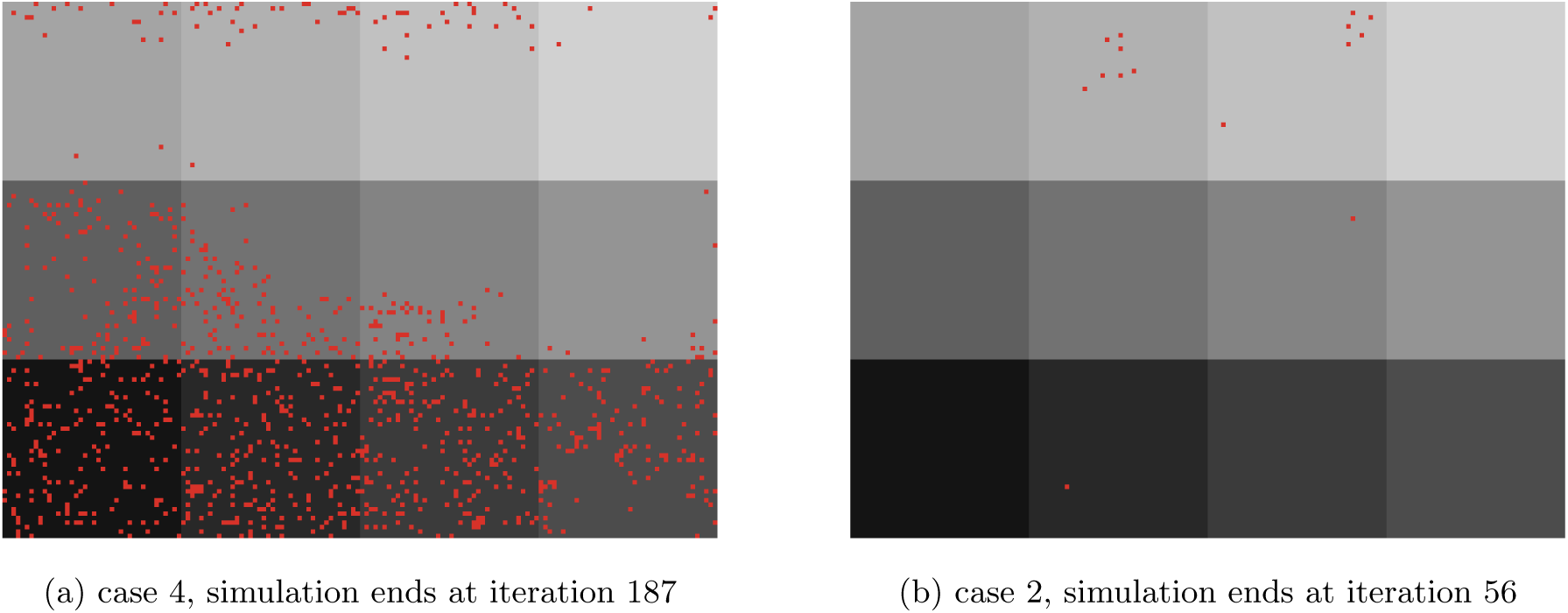
Regional Deaths: a red dots represent the place where at least one agent dies.

**Figure 8:**
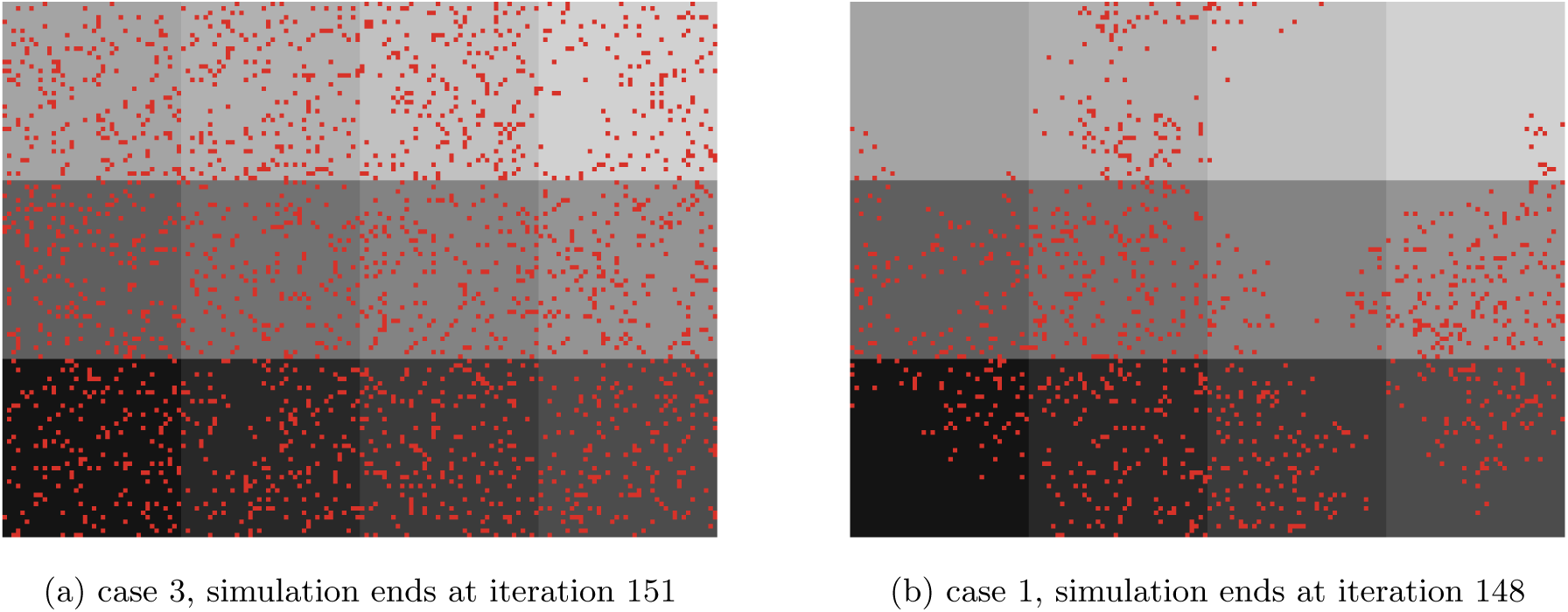
Regional Deaths: a red dots represent the place where at least one agent dies.

### dead?

It is clear from the graph shown in Figure 6 that a global vaccination drive, right from the start (in case of availability of vaccine) or whenever it was available, would have resulted in much less deaths. Now the question is do these deaths happen only in regions which are not self-centered. For this we consult the graph shown in Figure 9. As expected, in case 4, the number of people dead follow almost the same order from 1 to 12. But, if we compare it with case 2, case 2 gives a much improved global picture, in fact 10 times better than case 4. The gain among various blocks range from 5 times (for blocks with availability) to 15 times (for blocks with nonavailability). However, this disparity (between blocks) becomes really negligible if we compare case 3 with case 1. But, the global gain of case 1 is almost 2 times that of case 3. So, in future, in case of a pandemic (with availability of vaccine), it will be strategically beneficial to go for a global vaccination drive rather than the self-centered one.

**Figure 9:**
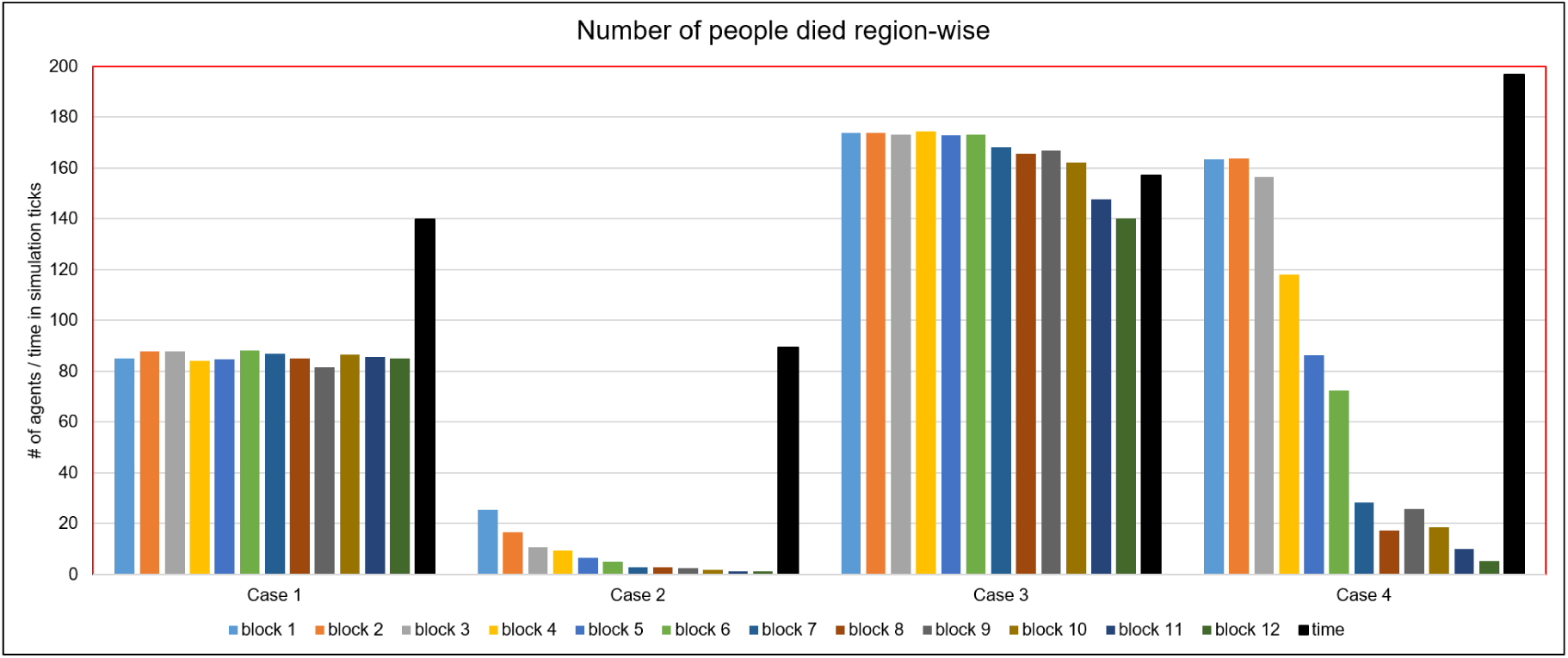
World arranged in vaccination blocks.

### time

A closure happening at a early time is absolutely beneficial as the World is tired and seeking a normal social and economic activity. At the time scale of the simulation, it is evident in the graph shown in Figure 9 that worst case time is for case 4. We gain at least 20% in case 2. When comparing case 3 with case 1, the gain is 10%. So global vaccination drive is also beneficial in terms of time needed to eradicate the pandemic. The timing dynamics aspect is visually shown in Figure 10 where we compare case 3 with case 1 in terms of time to vaccinate 20% of the population. In the sample run, it took 110 iterations to do it in case 3 as compared to 45 iterations in case 1.

**Figure 10:**
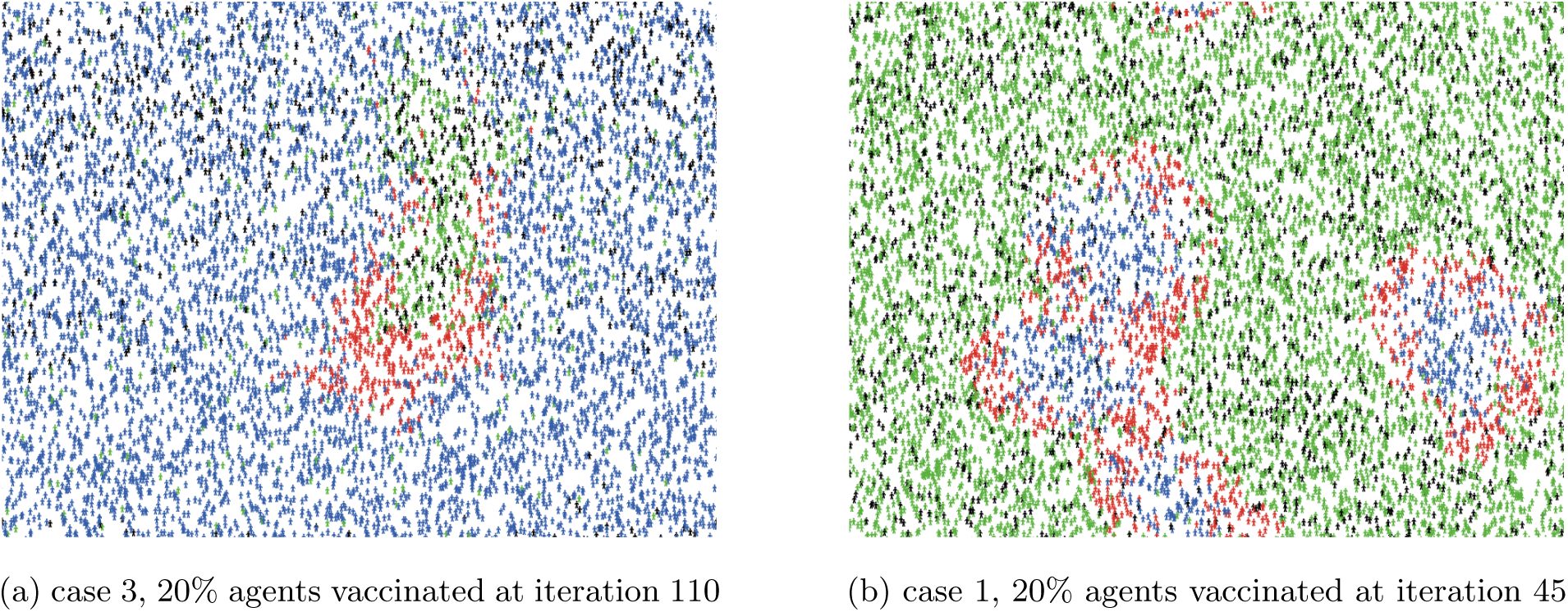
Timing: red = infected, blue = recovered, green = susceptible, and black = vaccinated.

### vaccinated?

Finally, we have results of how much population is needed to be vaccinated in each case in Figure 11. Obviously, if the vaccine is invented late (case 2 and 4), it will be required more, almost 85% of the population. And, there is no difference between these two cases. However, the time to achieve this number is lesser in case 4. Whereas, the time to achieve the required percentage between cases representing availability of vaccine right from the start is not much different among case 3 and 1, but, the required % decreases to almost 30% in case 3 when compared to more than 60% in case of case 1.

**Figure 11:**
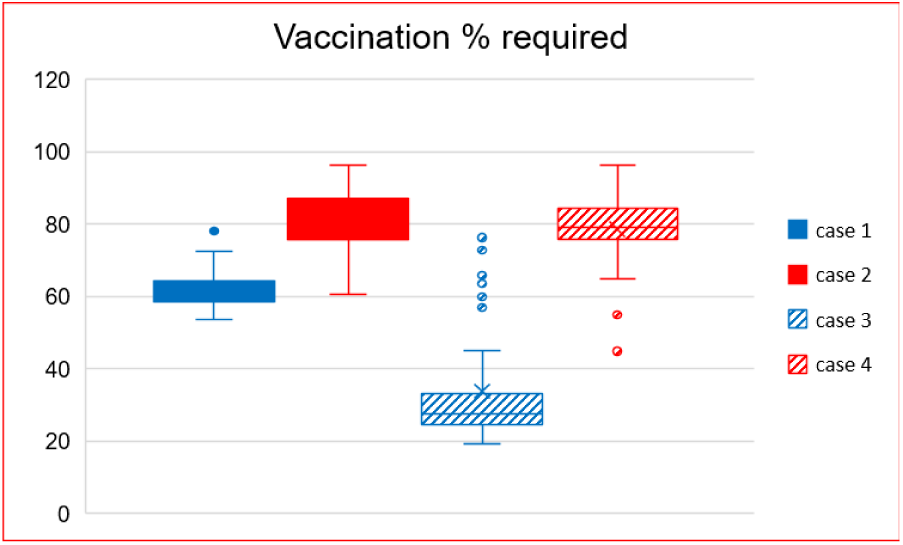
Vaccination percentage required.

## 4. Conclusion

There are several challenges in fair distribution of COVID-19 vaccine [12, 13]. In this paper, an Agent-Based Model is simulated to evaluate the effectiveness of COVID-19 vaccination drive based on its provision to different regions of the world. The model proposed is simple yet novel in the sense that it captures the spatial transmission-induced activity into consideration, through which we are able to relate transmission model to the mutated variations of the virus.

The results of the simulation suggest that it is necessary to maintain a global (rather than regional or country oriented) vaccination drive in case of a new pandemic or continual efforts against COVID-19. It results in lesser number of deaths, time and quantity of vaccination required.

The model can be extended in following ways:

- A more sophisticated virus transmission model can be used.
- The relationship of mortality and efficacy is dependent on variant of the virus. We have taken only static values, which can be replaced by evidenced values of mortality and efficacy based on the infectiousness of the virus.
- Real demographic, vaccination, population and mobility data can be used to gain a more insightful analysis.

## Data Availability

No data was used.

## Notes

### Competing Interest Statement

The authors have declared no competing interest.

### Funding Statement

No funding was used.

